# Integrated serosurveillance to assess disease elimination in coastal Ecuador: onchocerciasis, yaws, and trachoma

**DOI:** 10.64898/2026.02.16.26346420

**Authors:** Lesly Simbaña Vivanco, Stuart Torres Ayala, Nikolina Walas, Gretchen Cooley, Chabier Coleman, E. Brook Goodhew, Diana L. Martin, Hadley Burroughs, Everlyn Kamau, Manuel Calvopiña, William Cevallos, Josefina Coloma, Gwenyth O. Lee, Gabriel Trueba, Joseph N.S. Eisenberg, Karen Levy, Benjamin F. Arnold

## Abstract

**Background:** Testing blood samples with multiplex bead assays can assess elimination and identify residual foci of transmission for multiple pathogens simultaneously. In Ecuador, onchocerciasis and yaws are presumed eliminated, and the status of trachoma is unknown. We assessed their elimination status by measuring IgG antibodies in children 6–24 months.

**Methods:** In a birth cohort of 404 children measured between 2021-2024 in Esmeraldas province, Ecuador, we tested dried blood spots at ages 6, 9, 12, 18, and 24 months using a multiplex IgG assay that included antigens to *Onchocerca volvulus* (Ov16), *Treponema pallidum* (rp17, tmpa), and *Chlamydia trachomatis* (Pgp3, Ct694). We estimated seroprevalence and incident seroconversion rates across an urban-rural gradient.

**Results:** Of 404 children enrolled, 370 contributed 1,606 samples. Seroprevalence was near zero for onchocerciasis (0.4%, 95% CI: 0.2% to 0.8%) and yaws (0.2%, 95% CI: 0.0% to 0.5%). Conversely, *C. trachomatis* Pgp3 seroprevalence increased along the urban-rural gradient from Esmeraldas city (3.4%, 95% CI: 1.8% to 5.8%) to remote, river-accessible rural villages (22.4%, 95% CI: 16.0% to 30.1%), and incident seroconversion was common in rural villages (16.1 per 100 child-years, 95% CI: 11.3 to 21.9).

**Conclusions:** Serologic surveillance found no evidence of *O. volvulus* or *T. pallidum* transmission, consistent with elimination. High Pgp3 seroconversion rates suggest ongoing *C. trachomatis* transmission in rural villages. Results highlight the value of integrated serologic surveillance and motivate in-depth trachoma surveillance in Esmeraldas.

**Lay Summary:** Antibodies measured in blood provide a sensitive measure of past infection. At the population-level, antibody responses are widely used to monitor pathogen elimination. Antibodies measured in blood samples from children along an urban-rural gradient in Ecuador showed no evidence of exposure to the pathogens that cause onchocerciasis or yaws, confirming their continued elimination in the region. However, antibody responses to chlamydia were common among children in rural villages during their first two years, motivating additional surveillance to assess trachoma endemicity in the area as its endemicity status is currently unknown. Trachoma is caused by ocular chlamydia infection, and these results indicate a need for additional surveillance to assess ocular infections and clinical signs of trachoma in this population.

## Background

Multiplex antibody assays enable quantification of Immunoglobulin G (IgG) antibody responses to dozens of antigens from a single dried blood spot. Serological surveillance of routinely collected specimens using multiplex assays can support diverse public health priorities, such as monitoring pathogen elimination, pathogen emergence, or immunization gaps [1,2]. In recent years, a joint initiative between the Pan American Health Organization (PAHO), the World Health Organization, and the United States Centers for Disease Control and Prevention developed a framework for integrated serological surveillance in the Americas and identified monitoring disease elimination and pathogen reintroduction or reemergence as one of three key use cases for integrated platforms [3].

Esmeraldas province in coastal Ecuador has been a focus of research into infectious disease emergence and elimination in the Americas for decades. In the 1980s, 11 endemic foci of onchocerciasis (*Onchocerca volvulus*) were identified in Esmeraldas after being introduced from Africa, with high levels of endemicity along the Cayapas and Santiago rivers [4]. Repeated mass distribution of ivermectin in endemic communities from 1991-2009 led to transmission interruption and the region was declared free of onchocerciasis [5,6]. Epidemiologic surveys in the 1980s also revealed ongoing transmission of yaws (*Treponema pallidum* subspecies *pertenue*) in communities in the Santiago basin, including the Cayapas, Santiago, and Onzole rivers [7]. After repeated rounds of mass antibiotic distribution, yaws was declared eliminated in 1998 [8,9]. The status of trachoma (caused by ocular *Chlamydia trachomatis* infection) in Ecuador is unknown, but it is thought that foci could remain in tropical remote communities [10]. In 2023, PAHO launched the initiative for elimination of trachoma in the Americas, through which new surveillance of trachoma in Ecuador is ongoing with priority surveillance in the Amazon [11].

IgG antibodies provide a sensitive measure of previous infection and are a key tool for assessing pathogen elimination because young children born after elimination will not have IgG antibody responses to the eliminated pathogen beyond the first few months of life when residual maternal antibody will still be present. Due to its sensitivity, pathogen-specific IgG played a central role in the documentation of transmission interruption and confirmation of continued elimination of onchocerciasis and yaws in Ecuador [5,6,9,12,13]. For trachoma, use of IgG responses to the Pgp3 antigen in young children provides information about transmission of ocular *C. trachomatis*, and methods are in place to use the Pgp3 seroconversion rate to assess the need for public health intervention [14,15].

Here, we report results from multiplex IgG testing in a birth cohort from 2021-2024 along an urban-rural gradient in Esmeraldas, Ecuador, including villages along Santiago and Onzole rivers. The cohort was enrolled to study enteric pathogen infections and the developing microbiome [16]. The multiplex assay included antigens to *O. volvulus*, *T. pallidum*, and *C. trachomatis*, providing a unique opportunity to assess elimination through integrated serological surveillance in a previously endemic population.

## Methods

### Study design, population and setting

ECoMiD is a longitudinal birth cohort study in Esmeraldas Province, on the northern coast of Ecuador [16]. The study enrolled pregnant mothers from 2019 through 2022 along an urban-rural gradient from urban Esmeraldas city (urban), Borbón (a commercial center for the region), and eight rural villages along Rio Santiago and Rio Onzole that were accessible by road or river (**Figure 1**). The primary objective of the cohort is to study enteric pathogen transmission and the microbiome. Children in the birth cohort were followed for their first two years of life, with biological samples collected every three months. Within the cohort, 404 children enrolled after the SARS-CoV-2 pandemic (from 2021 onwards) were eligible to participate in serological testing. The study region is predominantly lowland tropical rainforest and converted agricultural land that has undergone gradual development since road construction increased in the early 2000s, bringing increased connectivity between urban Esmeraldas city, the town of Borbón, and rural villages.

**Figure 1.**
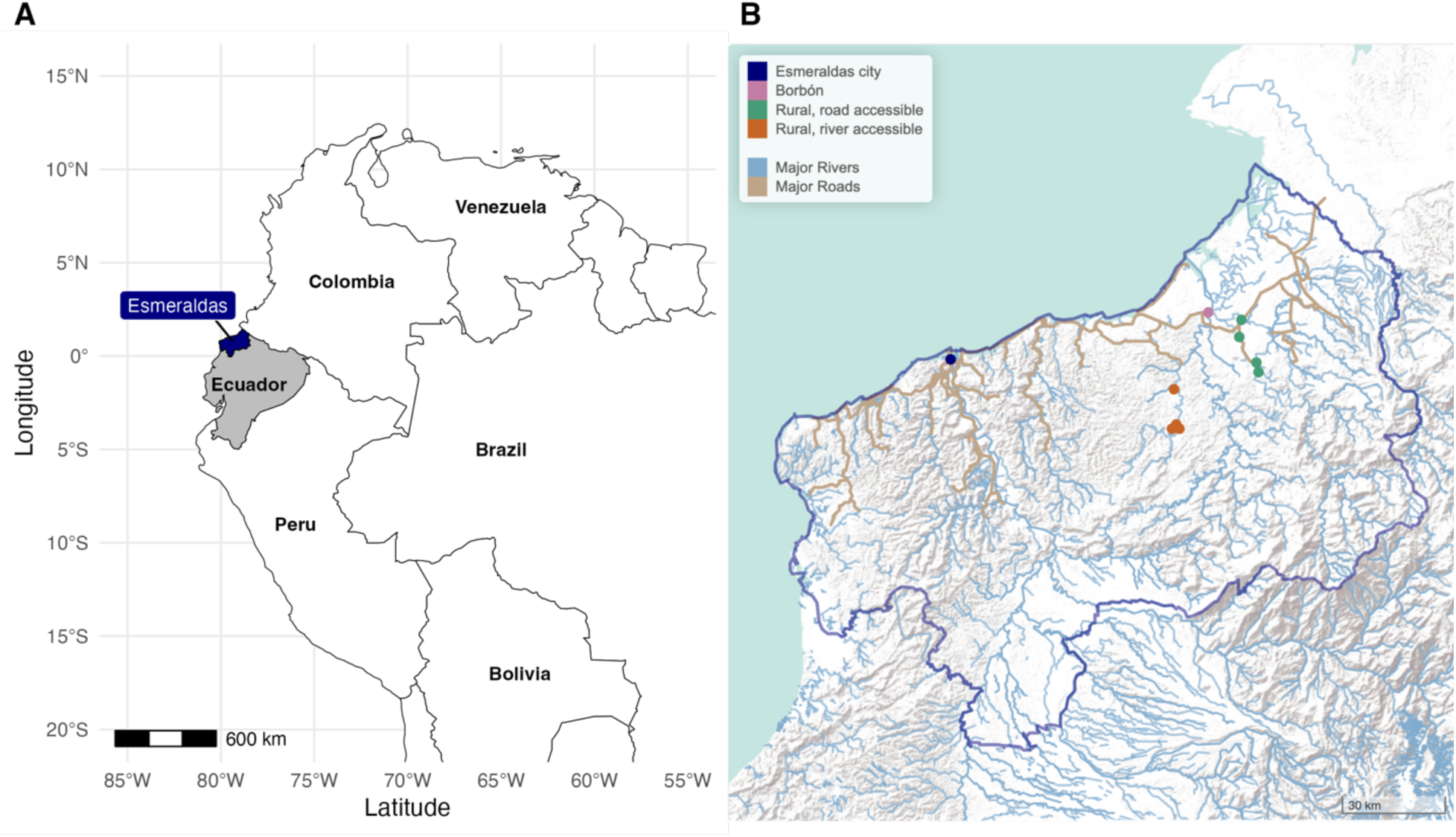
Study location of the ECoMiD birth cohort in Esmeraldas province, Ecuador. **A**. Esmeraldas province is located in northern coastal, Ecuador, along the border with Colombia. **B.** Children were enrolled from Esmeraldas city (population approximately 162,000), the town of Borbón (population approximately 5,000), four road accessible rural villages (populations 500 to 1000) and four river accessible rural villages (populations 200 to 700). Major rivers (blue) and roads (brown) provided by OpenStreetMap.

### Ethics

Institutional review boards at University of Washington (UW; STUDY00014270), Emory University (IRB00101202), University of California, San Francisco (21−33932), Universidad San Francisco de Quito (USFQ; 2018−022M), and the Ecuadorian Ministry of Health (MSPCURI000253−4) reviewed and approved the study protocol. Caregivers provided informed consent and field staff obtained consent and assent before each sample collection. Centers for Disease Control and Prevention (CDC) staff had no interaction with study participants and no access to identifying information so were determined to not be engaged in human subjects research.

### Sample collection and testing

Field staff collected dried blood spot samples from children up to five times at ages 6, 9, 12, 18, and 24 months. Dried blood spots were collected on filter paper, each sample provided six 10 μL spots. These were then dried, individually sealed with desiccant in plastic bags, and stored at - 20°C in the field office before being shipped to the USFQ at ambient temperature and stored at - 20°C.

A single 10 μL spot from each participant’s sample was eluted overnight at 4°C in a Buffer B (1X PBS pH 7.2-7.4, 0.5% casein, 0.5% PVA, 0.8% PVP, 0.3% Tween 20, 0.02% NaN3, 3 ug/ml of *E. coli* extract). Eluates were further diluted with Buffer B to a final concentration of 1:400 for use in the multiplex bead assay [17]. Eluted samples were incubated with a 45-plex of antigen-coupled beads to a broad set of enteric pathogens, vaccine preventable diseases, arboviruses, and neglected tropical diseases, including: Ov16 (*O. volvulus*) rp17 and TmpA (*T. pallidum*), and Pgp3 and Ct694 (*C. trachomatis*). The details of the sample preparation protocol have been published [17].

IgG bound to each antigen-coupled bead was detected using a Luminex MAGPIX® system (Luminex Corp., Austin, TX) with xMAP® Technology. Plates included Buffer B-only wells as blanks, along with a negative control and three pooled positive controls of known antigens at different concentrations for quality assurance. The presence of antigen-specific IgG was outputted as median fluorescence intensity with the Buffer B blank value subtracted from each antigen’s MFI values for a final ouput if MFI-background (MFI-bg). Plates were repeated if positive control monitoring targets fell outside two standard deviations of the mean in Levey-Jennings plots across all plates. Plate controls were run in duplicate to assess the coefficient of variation (CV) within the plate and across plates. A threshold of 20% CV for controls along with widespread low bead count (<10 beads) were used to flag plates for repeat analysis.

### Statistical analysis

Seropositivity cutoffs for Ov16 (198), rp17 (40), TmpA (154), Pgp3 (238), and Ct694 (710) were determined using external values from a receiver operator characteristic curve analysis of known positive and negative samples at the US CDC [18]. For *T. pallidum*, we classified samples as seropositive if MFI-bg values were above seropositivity cutoffs to both antigens [19]. For *C. trachomatis*, we focused primarily on Pgp3 responses to align with elimination surveillance studies that rely primarily on Pgp3, and used Ct694 as an adjunctive measure [14,15].

We estimated seroprevalence as the proportion of samples positive with exact binomial (Clopper-Pearson) 95% confidence intervals. For *C. trachomatis*, we further stratified seroprevalence estimates along an urban-rural gradient, from Esmeraldas city (urban), to Borbon, rural road-accessible villages and rural river-accessible villages. We estimated prevalence differences between the four groups (Esmeraldas city, Borbón, rural road, rural river) using a linear-binomial model with robust standard errors clustered at the child-level.

We further estimated prospective *C. trachomatis* seroconversion rates using longitudinal measurements. We assumed children were at risk if seronegative at the beginning of an interval and counted person-time at risk assuming that incident seroconversions occurred at the midpoint of the sampling interval. We similarly estimated seroreversion rates among children who had seroconverted [20–22]. Among children who were seropositive by Pgp3, we examined longitudinal trajectories of Ct694 as a confirmatory measure to rule out false positives, and re-estimated seroconversion rates requiring samples to be positive to both Pgp3 and Ct694 to increase specificity. We estimated 95% confidence intervals for rates with a non-parametric bootstrap that resampled children with replacement. In strata with fewer than four events, we estimated exact Poisson (Garwood) confidence intervals.

### Data availability

We used R statistical software (version 4.5.1) for analyses. Replication files for the analyses are available through the Open Science Framework (https://osf.io/njkta). Due to the small population of the study region, UCSF IRB determined that individual-level data, even if de-identified, would be considered potentially identifiable. Please contact the corresponding author regarding possible access to data underlying this analysis, pending appropriate IRB approval.

## Results

Of the 404 children, 370 (92%) contributed 1,630 blood samples, of which 1,606 (98.5%) passed quality control and contributed to the analysis (**Figure 2**). Household and maternal characteristics varied across the urban-rural gradient (**Table 1**).

**Figure 2.**
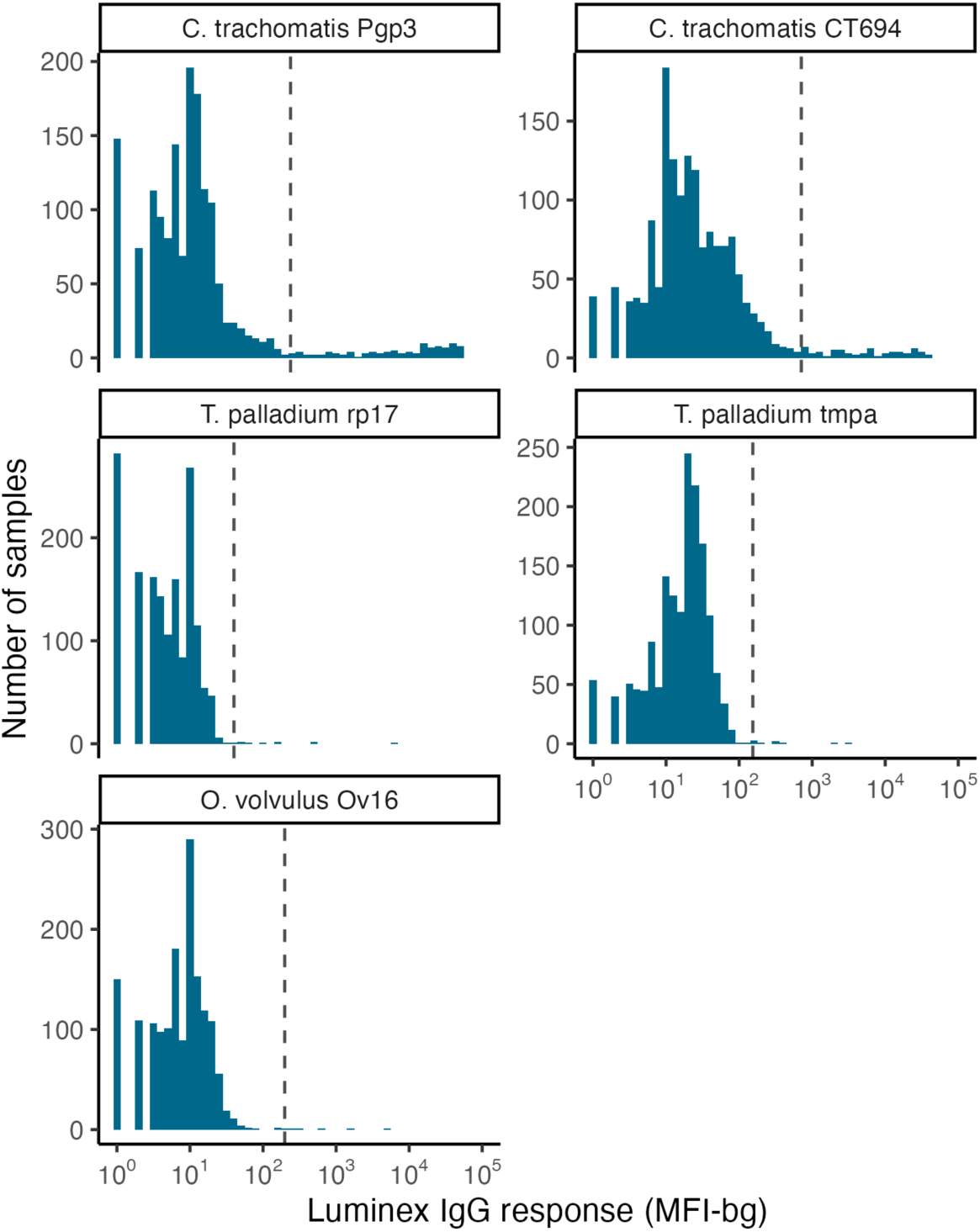
Distribution of IgG values by antigen among children aged 6–24 months in Esmeraldas province, Ecuador, 2021-2024. IgG was measured on the Luminex platform using Median Fluorescence Intensity minus background (MFI-bg). N=1,1605 measurements from 370 children. Vertical dashed lines represent seropositivity cutoffs.

**Table 1.**
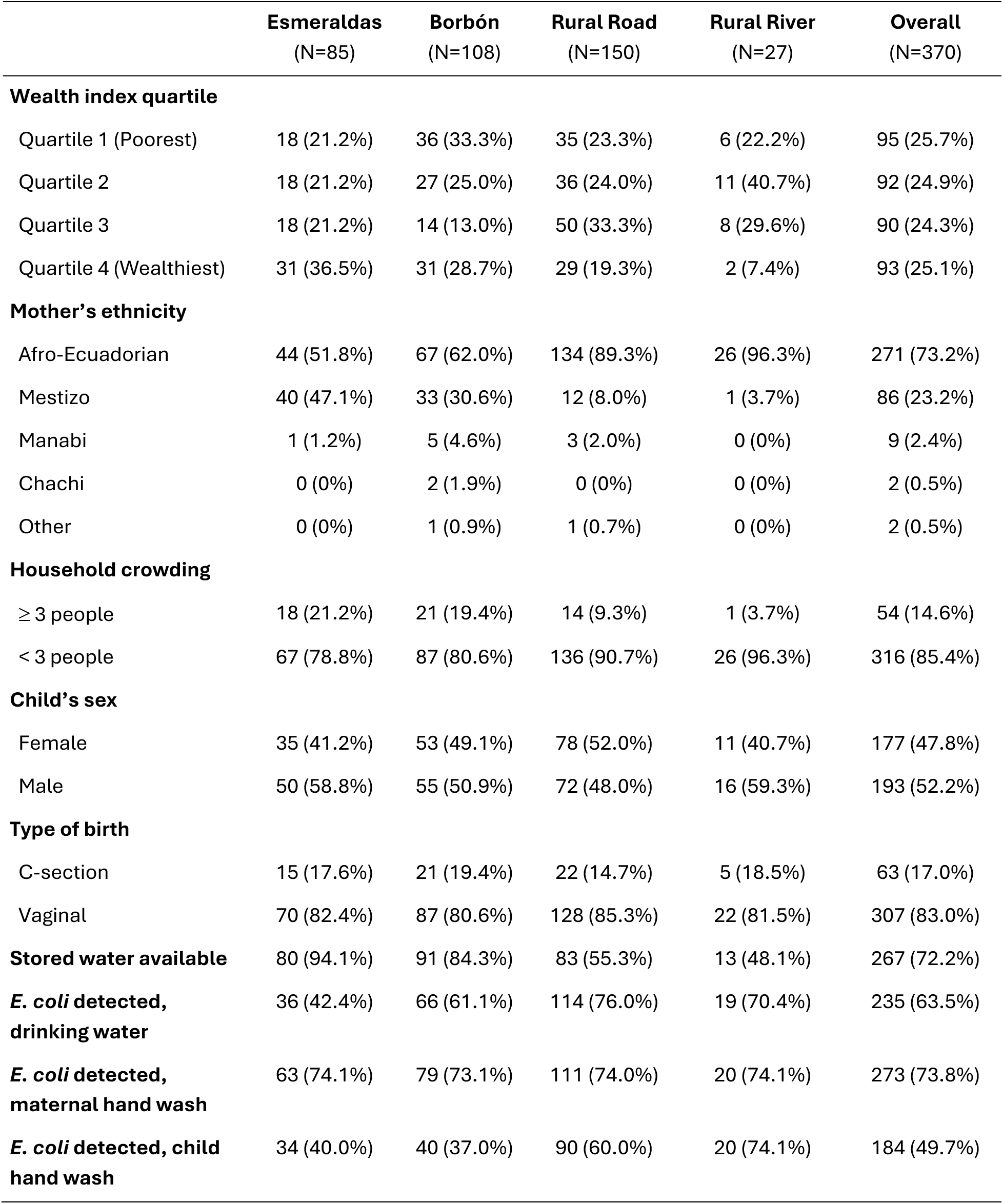
Characteristics of 370 children enrolled in the birth cohort along an urban-rural gradient. Esmeraldas city is an urban environment, Borbon is intermediate remoteness, and rural road-accessible and river-accessible communities are increasingly remote.

|  | Esmeraldas<br>(N=85) | Borbón<br>(N=108) | Rural Road<br>(N=150) | Rural River<br>(N=27) | Overall<br>(N=370) |
| --- | --- | --- | --- | --- | --- |
| **Wealth index quartile** |  |  |  |  |  |
| Quartile 1 (Poorest) | 18 (21.2%) | 36 (33.3%) | 35 (23.3%) | 6 (22.2%) | 95 (25.7%) |
| Quartile 2 | 18 (21.2%) | 27 (25.0%) | 36 (24.0%) | 11 (40.7%) | 92 (24.9%) |
| Quartile 3 | 18 (21.2%) | 14 (13.0%) | 50 (33.3%) | 8 (29.6%) | 90 (24.3%) |
| Quartile 4 (Wealthiest) | 31 (36.5%) | 31 (28.7%) | 29 (19.3%) | 2 (7.4%) | 93 (25.1%) |
| **Mother's ethnicity** |  |  |  |  |  |
| Afro-Ecuadorian | 44 (51.8%) | 67 (62.0%) | 134 (89.3%) | 26 (96.3%) | 271 (73.2%) |
| Mestizo | 40 (47.1%) | 33 (30.6%) | 12 (8.0%) | 1 (3.7%) | 86 (23.2%) |
| Manabi | 1 (1.2%) | 5 (4.6%) | 3 (2.0%) | 0 (0%) | 9 (2.4%) |
| Chachi | 0 (0%) | 2 (1.9%) | 0 (0%) | 0 (0%) | 2 (0.5%) |
| Other | 0 (0%) | 1 (0.9%) | 1 (0.7%) | 0 (0%) | 2 (0.5%) |
| **Household crowding** |  |  |  |  |  |
| ≥ 3 people | 18 (21.2%) | 21 (19.4%) | 14 (9.3%) | 1 (3.7%) | 54 (14.6%) |
| < 3 people | 67 (78.8%) | 87 (80.6%) | 136 (90.7%) | 26 (96.3%) | 316 (85.4%) |
| **Child's sex** |  |  |  |  |  |
| Female | 35 (41.2%) | 53 (49.1%) | 78 (52.0%) | 11 (40.7%) | 177 (47.8%) |
| Male | 50 (58.8%) | 55 (50.9%) | 72 (48.0%) | 16 (59.3%) | 193 (52.2%) |
| **Type of birth** |  |  |  |  |  |
| C-section | 15 (17.6%) | 21 (19.4%) | 22 (14.7%) | 5 (18.5%) | 63 (17.0%) |
| Vaginal | 70 (82.4%) | 87 (80.6%) | 128 (85.3%) | 22 (81.5%) | 307 (83.0%) |
| **Stored water available** | 80 (94.1%) | 91 (84.3%) | 83 (55.3%) | 13 (48.1%) | 267 (72.2%) |
| ***E. coli* detected, drinking water** | 36 (42.4%) | 66 (61.1%) | 114 (76.0%) | 19 (70.4%) | 235 (63.5%) |
| ***E. coli* detected, maternal hand wash** | 63 (74.1%) | 79 (73.1%) | 111 (74.0%) | 20 (74.1%) | 273 (73.8%) |
| ***E. coli* detected, child hand wash** | 34 (40.0%) | 40 (37.0%) | 90 (60.0%) | 20 (74.1%) | 184 (49.7%) |

The upper 95% confidence interval for seroprevalence to onchocerciasis and yaws antigens fell below 1%, consistent with no ongoing transmission in the study populations (**Table 2**). Positive samples to onchocerciasis and yaws likely represented false positives since assay specificity is not 100%.

**Table 2.** Seroprevalence to three pathogens assessed for elimination among 370 children measured between ages 6 and 24 months in Esmeraldas province, Ecuador 2021-2024.

| Disease, pathogen (antigens) | N<br>Tested* | N<br>Positive | Seroprevalence<br>(95% CI) |
| --- | --- | --- | --- |
| Trachoma, *C. trachomatis* (Pgp3) | 1,601 | 104 | 6.5 (5.3 to 7.8) |
| Onchocerciasis, *O. volvulus* (Ov16) | 1,605 | 6 | 0.4 (0.1 to 0.8) |
| Yaws, *T. pallidum* (rp17, tmpa) | 1,605 | 3 | 0.2 (0.0 to 0.5) |

\* Of 1,606 dried blood spot samples collected from 370 children, some targets failed on the Luminex platform or did not pass quality control, which is why the number tested varies across pathogens.

In contrast, 104 samples (6.5%) were positive to *C. trachomatis* Pgp3, and seroprevalence increased along the urban-rural gradient (**Table 3**). Due to relatively small sample size in rural river-accessible villages, we combined all eight rural villages into a single group to study incident seroconversions. Seroconversion rates to Pgp3 were consistent with postnatal *C. trachomatis* transmission in rural villages, even after a more stringent definition that required a child seroconvert to both Pgp3 and Ct694 antigens (**Table 4**). Cumulative incidence of seroconversion recapitulated results based on the incidence rate, with 19.4% of children in rural villages seroconverting between ages 6-24 months (**Supplementary Table 1**).

**Table 3.** *C. trachomatis* Pgp3 seroprevalence by community type among 370 children measured between ages 6-24 months in Esmeraldas province, Ecuador 2021-2024.

| Location | N<br>Tested | N<br>Positive | Seroprevalence<br>(95% CI) | Difference*<br>(95% CI) | P-value |
| --- | --- | --- | --- | --- | --- |
| Esmeraldas city | 355 | 12 | 3.4 (1.8 to 5.8) | Ref. |  |
| Borbón | 468 | 17 | 3.6 (2.1 to 5.8) | 0.3 (−2.3 to 2.8) | 0.85 |
| Rural, road accessible | 631 | 42 | 6.7 (4.8 to 8.9) | 3.3 (0.6 to 6.0) | 0.02 |
| Rural, river accessible | 147 | 33 | 22.4 (16.0 to 30.1) | 19.1 (12.1 to 26.1) | <0.001 |

\* Difference in prevalence estimated using linear binomial models with robust standard errors clustered at the child level

**Table 4.** *Chlamydia trachomatis* IgG seroconversion rates (SCR) estimated longitudinally among children ages 6-24 months in Esmeraldas Province, Ecuador 2021-2024. Estimates are presented for seroconversion to Pgp3 alone and for seroconversion to both Pgp3 and Ct694 antigens. Rural villages include eight communities accessed by road or river.

| Population | N children | Incident seroconversions | Person-years at risk | Seroconversion Rate (95% CI) * |
| --- | --- | --- | --- | --- |
| *Pgp3 alone* |  |  |  |  |
| **Overall** | 370 | 38 | 431.9 | 8.8 (6.3 to 11.7) |
| **Location** |  |  |  |  |
| Esmeraldas city | 85 | 4 | 98.8 | 4.0 (0.0 to 9.9) |
| Borbón | 108 | 2 | 134.1 | 1.5 (0.2 to 5.4) |
| Rural villages | 177 | 32 | 199.0 | 16.1 (11.3 to 21.9) |
| *Pgp3 & Ct694* |  |  |  |  |
| **Overall** | 370 | 21 | 445.8 | 4.7 (2.7 to 7.1) |
| **Location** |  |  |  |  |
| Esmeraldas city | 85 | 1 | 100.4 | 1.0 (0.0 to 5.5) |
| Borbón | 108 | 1 | 136.5 | 0.7 (0.0 to 4.1) |
| Rural villages | 177 | 19 | 298.9 | 9.1 (5.2 to 14.2) |

\* SCR: seroconversion rate per 100 person-years. SCR was estimated longitudinally with bootstrap 95% confidence intervals that resampled children with replacement. For strata with fewer than 4 events the 95% CI was estimated using exact Poisson (Garwood) method.

Given this finding, we conducted a series of additional analyses to assess the dynamics of Pgp3 serology in the cohort. Longitudinal trajectories showed maternal IgG decay for some children from 6-9 months, while others sustained high IgG from age 6 months onward, potentially from congenital infection (**Supplementary Figure 1**). Of 18 children who were seropositive at 6 months (4.9%), all were born by vaginal delivery — although not necessarily a causal relationship, the observation is consistent with congenital infection (**Supplementary Table 2**).

Among children who seroconverted after 6 months, most lost IgG rapidly in the following months (**Supplementary Figure 1**). The Pgp3 seroreversion rate in the 12-24-month period, excluding potential maternal IgG, was 92.1 per 100 child-years (**Supplementary Table 3**). Concordance between Pgp3 and Ct694 IgG trajectories among children who seroconverted to both antigens (31 children, **Supplementary Figure 2**) and even among those who seroconverted only to Pgp3 (24 children, **Supplementary Figure 3**) suggest false positives were unlikely.

## Discussion

This analysis of IgG responses among young children in coastal Ecuador demonstrates the value of integrated serologic surveillance to inform several disease elimination programs. Antibody measurements in young children provide a sensitive measure of past infection and were consistent with elimination of onchocerciasis and yaws in a previously endemic population. High seroprevalence and incident seroconversion to Pgp3 between ages 6 and 24 months in rural villages is suggestive of ongoing transmission of *C. trachomatis* among children.

Little is known about the status of trachoma elimination in Ecuador [3,11]. The only epidemiologic study of *C. trachomatis* we could identify from the region was a 1989 report that documented high prevalence of *C. trachomatis* among women in Borbón [23]. As we approach the goal of global trachoma elimination as a public health problem by 2030, broader testing within existing serological surveys can help identify previously unknown areas of endemicity [3,15]. Our results motivate additional follow-up to measure ocular chlamydia infection, clinical signs of trachoma, and, if warranted, public health intervention.

The riverine communities included in this study have abundant access to surface water unlike many current or previously trachoma-endemic populations in Ethiopia, Sudan, or Niger. Yet, a recent survey in Alto Amazonas, Peru estimated trachoma prevalence of >25% in rural villages situated in a similar tropical, riparian biome [24], suggesting that the availability of water for face washing is potentially insufficient on its own to prevent *C. trachomatis* transmission among young children in remote, tropical areas of South America. A high burden of enteric pathogen infection in the ECoMiD birth cohort [25] is also consistent with conditions that facilitate person-to-person bacterial pathogen transmission to infants.

The study had limitations. Opportunistic testing of samples in a birth cohort meant that the sampling design does not align with designs recommended to verify elimination for onchocerciasis, yaws, and trachoma [26–28]. So while not definitive, results for onchocerciasis, and yaws help reaffirm official verification status by PAHO, further reinforcing recent, opportunistic serological testing of banked specimens in the region that measured *O. volvulus* Ov16 IgG and *T. pallidum* using rapid antibody tests [12,13]. The cohort included nearly all children born in rural villages during the study period, but the total number of children tested was relatively small and only 177 children lived in the rural regions known to be previously endemic for onchocerciasis and yaws. For the analysis of incident seroconversions to Pgp3, we combined data across all rural villages due to relatively rare events, likely averaging over important heterogeneity as prevalence measures suggested higher exposure in river-accessible villages. Nevertheless, high rates of seroconversion to *C. trachomatis* in rural villages provides a robust indicator to motivate additional follow-up.

Despite these limitations, the study had important strengths, including enrollment across an urban-rural gradient to capture heterogeneity in transmission combined with longitudinal measurements to identify incident seroconversion and seroreversion rates. A unique contribution of the longitudinal birth cohort was characterization of Pgp3 IgG dynamics from ages 6 to 24 months. Pgp3 IgG responses were transient, with frequent and rapid seroreversion. Absence of a robust, persistent IgG response among infants might be expected given the nascent development of infants’ humoral immunity, particularly for mucosal infections [29], but to our knowledge there are no previous reports of Pgp3 IgG dynamics in infants. Pgp3 seroreversion rates among older children ages 1-9 years in endemic settings have been lower than those estimated in this cohort [20–22]. Pgp3 seroconversion rate estimates from cross-sectional surveys include children 1-5 years old [15], so it is unclear if the high rates of seroreversion observed in 1-2 year olds would sustain at older ages.

In conclusion, multiplex antibody testing within an ongoing birth cohort found no antibody responses to onchocerciasis and yaws, consistent with continued absence of transmission in Esmeraldas province, Ecuador. High levels of seroprevalence and incident seroconversion to *C. trachomatis* Pgp3 among children 6 to 24 months old provide new information about possible trachoma endemicity in remote, rural communities and motivate additional trachoma surveillance in the region. The study demonstrates the value of integrated serosurveillance to inform public health elimination programs.

## Acknowledgements

We are grateful to the project’s dedicated local field team who completed all household visits and collected samples: Mauricio Ayovi, Adriana Lupero, Gabriela Vasco, Diana Barreiro, Mariuxi Caicedo, Grace Macias, Maria Nela Obando, Karen Caicedo, Yesica Perlaza, Yina Segura, Jorge Mejia, Juleisy Palma, Ana Rosero, Tamara Valencia, Zaida Jimenez, Sara Charcopa, Dasny Monrroy, Denise Angulo, Naira Martinez, and Dayanara Wila, and to the families who participate in the ECoMiD birth cohort. Author contributions, following CRediT taxonomy, conceptualization (LSV, BFA), data curation (LSV, STA, NW), formal analysis (LSV, BFA), funding acquisition (GT, JNSE, KL, BFA), investigation (LSV, STA, NW, GC, CC, EBG, HB), methodology (LSV, NW, EK, BFA), project administration (LSV, STA, GC, CC, WC, JC, GOL, GT, JNSE, KL, BFA), software (LSV, NW, BFA), supervision (DLM, GOL, GT, JNSE, KL, BFA), validation (NW, GC, CC, EBG), visualization (LSV, NW, BFA), Writing—Original Draft Preparation (LSV, BFA), Writing—Review & Editing (LSV, STA, NW, GC, CC, EBG, DLM, HB, EK, MC, WC, JC, GOL, GT, JNSE, KL, BFA).

## Supplementary Information Materials

**Supplementary Table 1.**
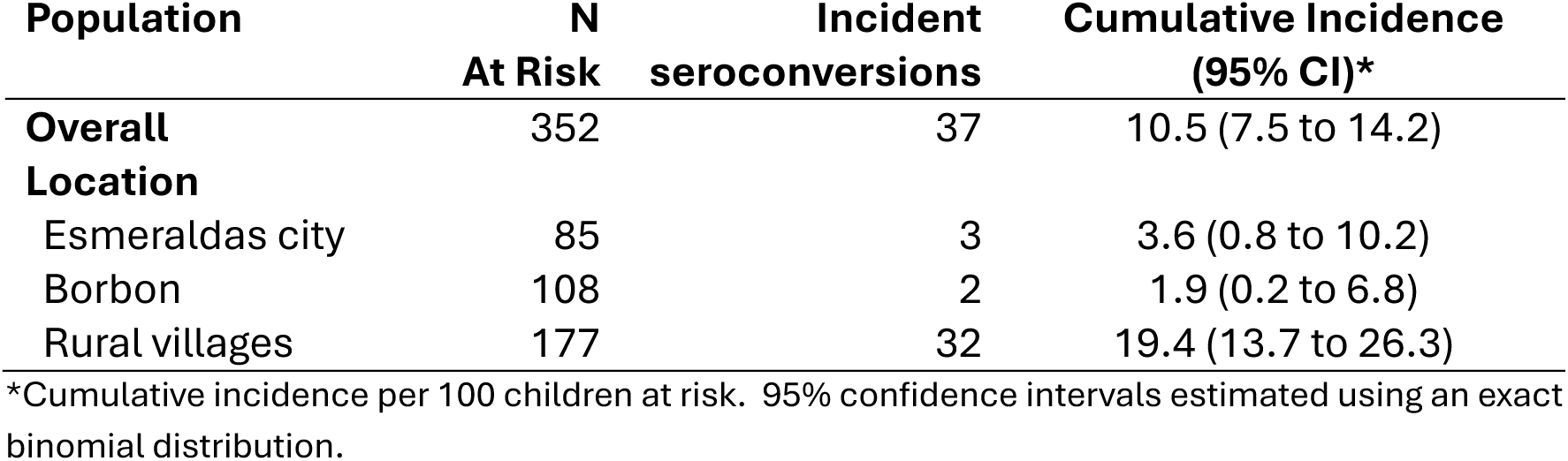
Cumulative incidence of *Chlamydia trachomatis* IgG seroconversion between ages 6 and 24 months in Esmeraldas Province, Ecuador 2021-2024. Esmeraldas is an urban environment, Borbon is intermediate urbanicity, and Rural villages include eight communities accessed by road or river.

| Population | N<br>At Risk | Incident<br>seroconversions | Cumulative Incidence<br>(95% CI)* |
| --- | --- | --- | --- |
| **Overall** | 352 | 37 | 10.5 (7.5 to 14.2) |
| **Location** |  |  |  |
| Esmeraldas city | 85 | 3 | 3.6 (0.8 to 10.2) |
| Borbon | 108 | 2 | 1.9 (0.2 to 6.8) |
| Rural villages | 177 | 32 | 19.4 (13.7 to 26.3) |

\*Cumulative incidence per 100 children at risk. 95% confidence intervals estimated using an exact binomial distribution.

**Supplementary Table 2.** Seroprevalence to *Chlamydia trachomatis* Pgp3 IgG at age 6 months in Esmeraldas province, Ecuador, 2021-2024. Esmeraldas is an urban environment, Borbon is intermediate urbanicity, and Rural villages include eight communities accessed by road or river.

| Population | N children | N positive | Seroprevalence<br>(95% CI)* |
| --- | --- | --- | --- |
| **Overall** | 370 | 18 | 4.9 (2.9 to 7.6) |
| **Birth type** |  |  |  |
| Cesarean section | 63 | 0 | 0.0 (0.0 to 5.7) |
| Vaginal delivery | 307 | 18 | 5.9 (3.5 to 9.1) |
| **Location** |  |  |  |
| Esmeraldas city | 85 | 2 | 2.4 (0.3 to 8.2) |
| Borbon | 108 | 4 | 3.7 (1.0 to 9.2) |
| Rural villages | 177 | 12 | 6.8 (3.6 to 11.5) |

\* 95% confidence intervals estimated using an exact binomial distribution.

**Supplementary Table 3.** *Chlamydia trachomatis* Pgp3 IgG seroreversion rates (SRR) estimated longitudinally among children ages 6-24 months in Esmeraldas Province, Ecuador 2021-2024. Rates were estimated over the whole age range, 6-24 months, and restricted to 12-24 months to avoid potential for seroreversion due to waning maternal IgG.

| Age range | N<br>children | Incident<br>sero-<br>reversions | Person-years at<br>risk | SRR<br>(95% CI) * |
| --- | --- | --- | --- | --- |
| 6 to 24 months | 370 | 26 | 23.3 | 111.4 (68.8 to 180.8) |
| 12 to 24 months | 352 | 13 | 14.1 | 92.1 (46.4 to 160.2) |

\* SRR: seroreversion rate per 100 person-years. SRR was estimated longitudinally with bootstrap 95% confidence intervals that resampled children with replacement.

**Supplementary Figure 1.**
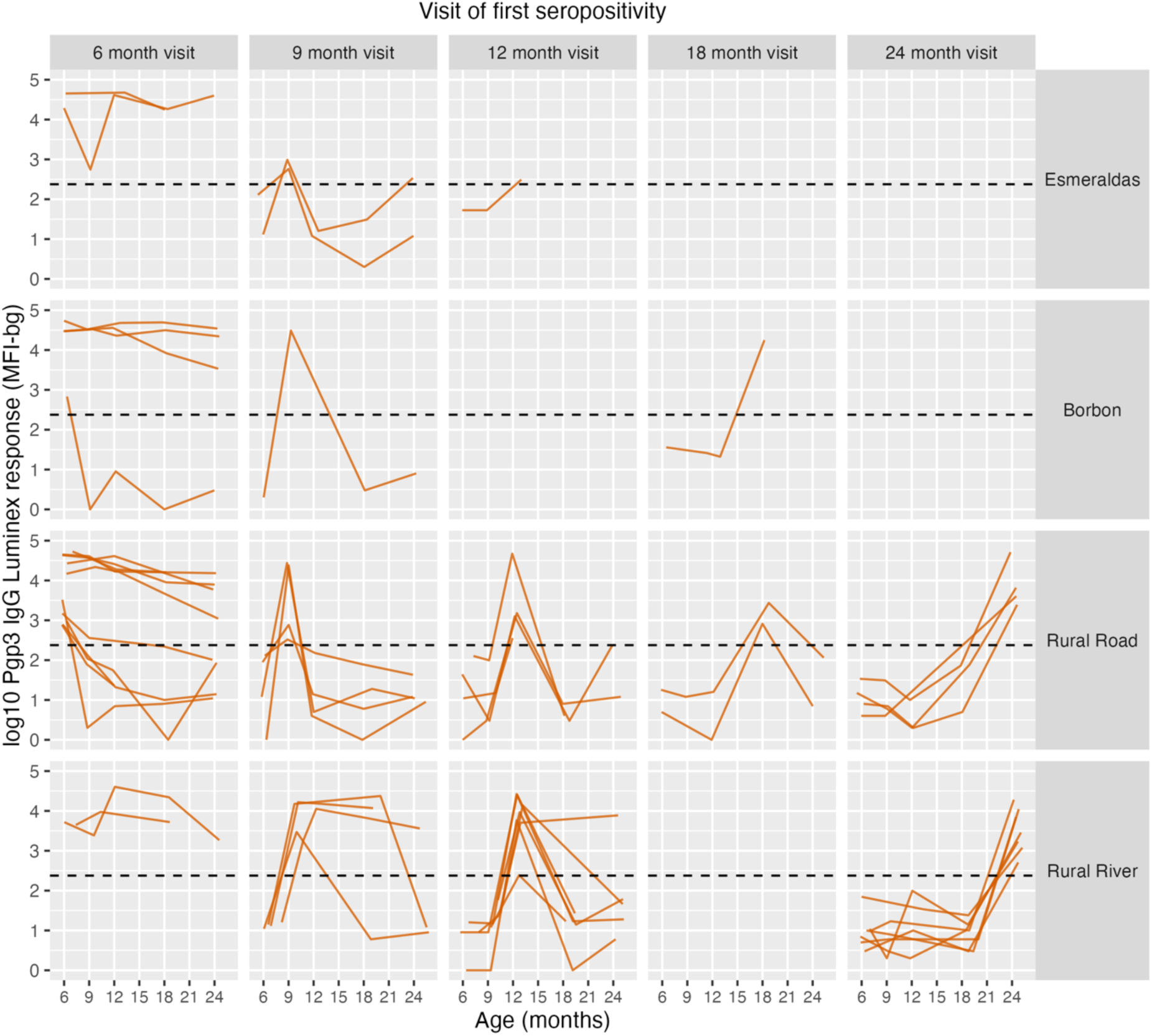
Longitudinal trajectories of *Chlamydia trachomatis* IgG responses to Pgp3 antigen among 55 children in the cohort who were seropositive during follow-up between ages 6 and 24 months in Esmeraldas, Ecuador, 2021-2024. Children are stratified by the visit at which they were first identified as seropositive to Pgp3 and by community type. IgG measured in Median Florescence Units minus background (MFI-bg) on the Luminex platform. A horizontal dashed line marks the seropositivity cutoR.

**Supplementary Figure 2.**
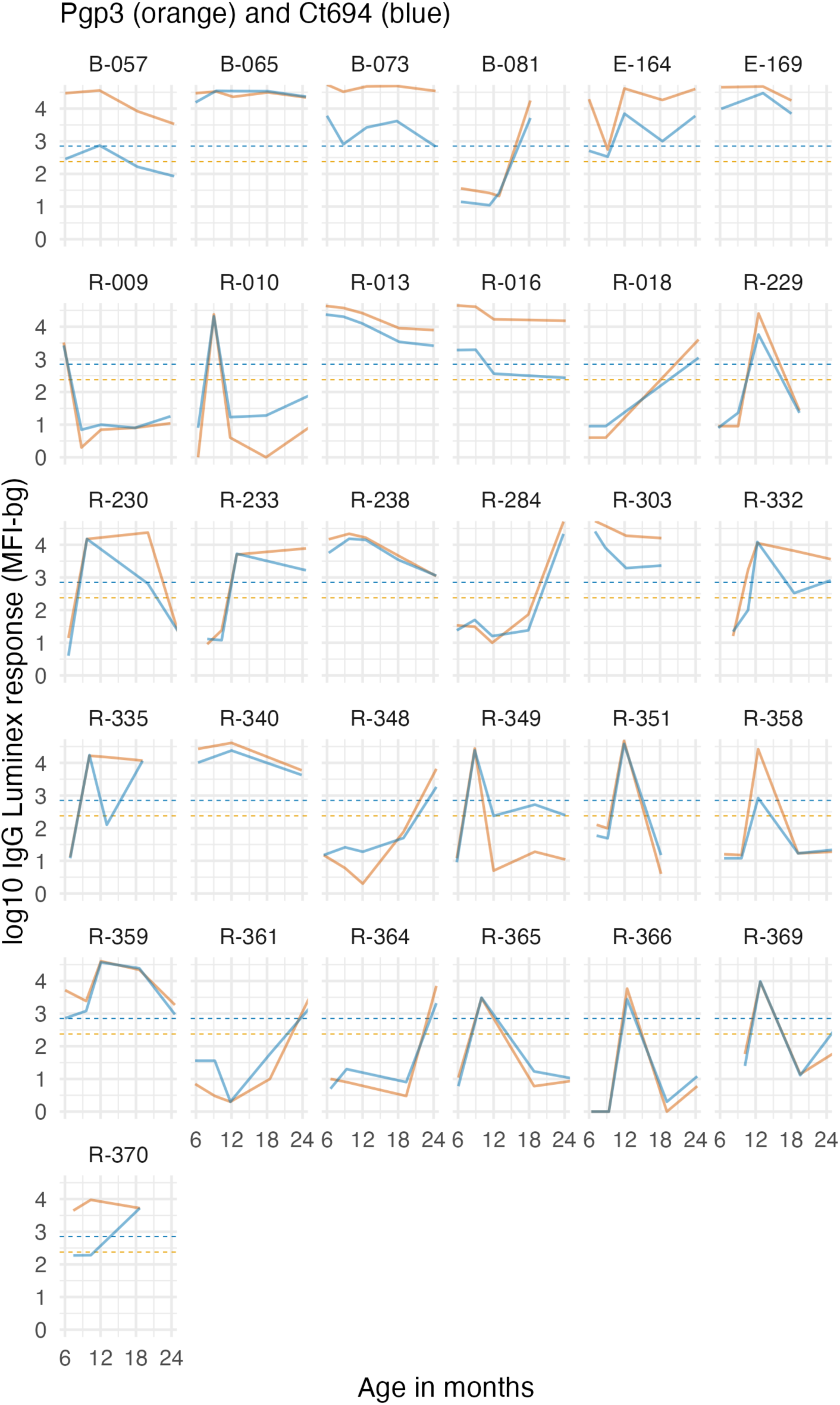
Longitudinal trajectories of *Chlamydia trachomatis* IgG responses to Pgp3 and Ct694 antigens among 31 children in the cohort who were seropositive to both antigens during follow-up between ages 6 and 24 months in Esmeraldas, Ecuador, 2021-2024. IgG measured in Median Florescence Units minus background (MFI-bg) on the Luminex platform.

**Supplementary Figure 3.**
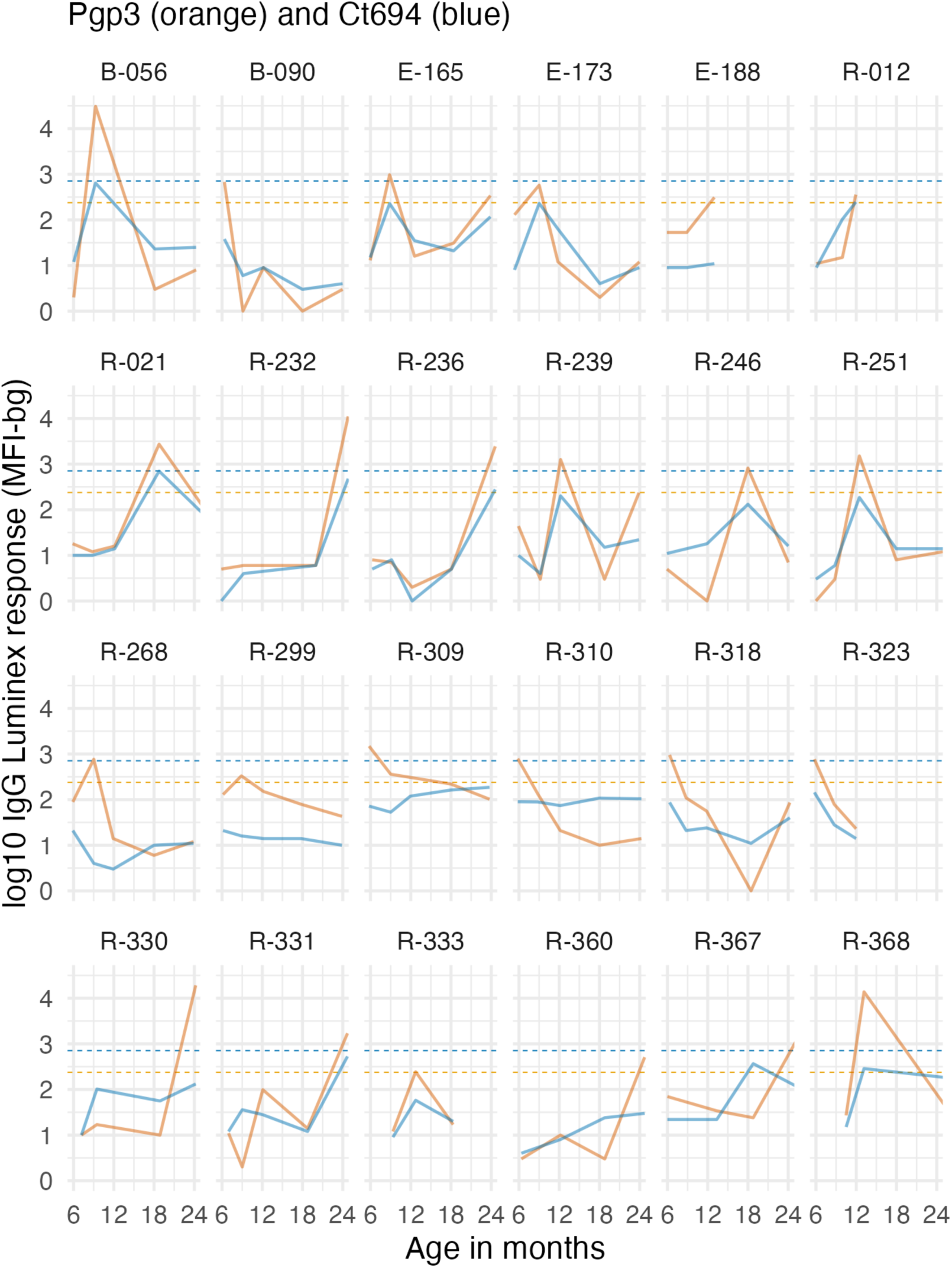
Longitudinal trajectories of *Chlamydia trachomatis* IgG responses to Pgp3 and Ct694 antigens among 24 children in the cohort who were seropositive to Pgp3 alone (not Ct694) during follow-up between ages 6 and 24 months in Esmeraldas, Ecuador, 2021-2024. IgG measured in Median Florescence Units minus background (MFI-bg) on the Luminex platform.

## References

1. Arnold BF, Scobie HM, Priest JW, Lammie PJ. Integrated Serologic Surveillance of Population Immunity and Disease Transmission. Emerg Infect Dis 2018; 24:1188–1194.

2. Wiens KE, Jauregui B, Arnold BF, et al. Building an integrated serosurveillance platform to inform public health interventions: Insights from an experts’ meeting on serum biomarkers. PLoS Negl Trop Dis 2022; 16:e0010657.

3. PAHO/WHO. Toolkit for Integrated Serosurveillance of Communicable Diseases in the Americas. 2022. Available at: https://www.paho.org/en/documents/toolkit-integrated-serosurveillance-communicable-diseases-americas. Accessed 10 December 2025.

4. Guderian RH, Molea J, Swanson D, Proaño R, Carrillo R, Swanson WL. Onchocerciasis in Ecuador. I. Prevalence and distribution in the province of Esmeraldas. Tropenmed Parasitol 1983; 34:143–148.

5. Lovato R, Guevara A, Guderian R, et al. Interruption of Infection Transmission in the Onchocerciasis Focus of Ecuador Leading to the Cessation of Ivermectin Distribution. PLOS Neglected Tropical Diseases 2014; 8:e2821.

6. Guevara Á, Lovato R, Proaño R, et al. Elimination of onchocerciasis in Ecuador: findings of post-treatment surveillance. Parasit Vectors 2018; 11:265.

7. Guderian RH, Guzman JR, Calvopiña M, Cooper P. Studies on a focus of yaws in the Santiago Basin, province of Esmeraldas, Ecuador. Trop Geogr Med 1991; 43:142–147.

8. Anselmi M, Araujo E, Narváez A, Cooper PJ, Guderian RH. Yaws in Ecuador: impact of control measures on the disease in the Province of Esmeraldas. Genitourin Med 1995; 71:343–346.

9. Anselmi M, Moreira J-M, Caicedo C, Guderian R, Tognoni G. Community participation eliminates yaws in Ecuador. Trop Med Int Health 2003; 8:634–638.

10. Saboyá-Díaz MI, Betanzos-Reyes AF, West SK, Muñoz B, Castellanos LG, Espinal M. Trachoma elimination in Latin America: prioritization of municipalities for surveillance activities. Rev Panam Salud Publica 2019; 43:e93.

11. PAHO. Elimination of trachoma in the Americas, Ecuador factsheet. 2024: 2. Available at: https://www.paho.org/sites/default/files/2024-03/2024-cde-paho-canada-trachoma-public-health-ecuador.pdf. Accessed 10 December 2025.

12. Cooper PJ, Anselmi M, Caicedo C, et al. Yaws elimination in Ecuador: Findings of a serological survey of children in Esmeraldas province to evaluate interruption of transmission. PLoS Negl Trop Dis 2022; 16:e0010173.

13. Guevara Á, Salazar E, Vicuña Y, et al. Use of Ov16-Based Serology for Post-Elimination Surveillance of Onchocerciasis in Ecuador. Am J Trop Med Hyg 2020; 103:1569–1571.

14. Tedijanto C, Solomon AW, Martin DL, et al. Monitoring transmission intensity of trachoma with serology. Nat Commun 2023; 14:3269.

15. Kamau E, Ante-Testard PA, Gwyn S, et al. Characterizing trachoma elimination using serology. Nat Commun 2025; 16:5545.

16. Lee GO, Eisenberg JNS, Uruchima J, et al. Gut microbiome, enteric infections and child growth across a rural-urban gradient: protocol for the ECoMiD prospective cohort study. BMJ Open 2021; 11:e046241.

17. Priest JW, Moss DM. Measuring Cryptosporidium Serologic Responses by Multiplex Bead Assay. Methods Mol Biol 2020; 2052:61–85.

18. Chan Y, Fornace K, Wu L, et al. Determining seropositivity-A review of approaches to define population seroprevalence when using multiplex bead assays to assess burden of tropical diseases. PLoS Negl Trop Dis 2021; 15:e0009457.

19. Cooley GM, Mitja O, Goodhew B, et al. Evaluation of Multiplex-Based Antibody Testing for Use in Large-Scale Surveillance for Yaws: a Comparative Study. J Clin Microbiol 2016; 54:1321–1325.

20. West SK, Munoz B, Kaur H, et al. Longitudinal change in the serology of antibodies to Chlamydia trachomatis pgp3 in children residing in a trachoma area. Sci Rep 2018; 8:3520.

21. West SK, Munoz B, Mkocha H, Gaydos CA, Quinn TC. The effect of Mass Drug Administration for trachoma on antibodies to Chlamydia trachomatis pgp3 in children. Sci Rep 2020; 10:15225.

22. Tedijanto C, Aragie S, Gwyn S, et al. Seroreversion to Chlamydia trachomatis Pgp3 Antigen Among Children in a Hyperendemic Region of Amhara, Ethiopia. J Infect Dis 2024; 230:293–297.

23. Narváez M, López Jaramillo P, Guevara A, Izurieta A, Guderian R. [Prevalence of Chlamydia trachomatis and Neisseria gonorrhoeae in 3 groups of Ecuadorian women with different sexual behaviors]. Bol Oficina Sanit Panam 1989; 107:220–225.

24. Nesemann JM, Morocho-Alburqueque N, Quincho-Lopez A, et al. Discrepancy between active trachoma and tests of chlamydial infection in Alto Amazonas, Peru. Clin Infect Dis 2025; :ciaf506.

25. Jesser KJ, Zhou NA, Hemlock C, et al. Environmental Exposures Associated with Enteropathogen Infection in Six-Month-Old Children Enrolled in the ECoMiD Cohort along a Rural-Urban Gradient in Northern Ecuador†. Environ Sci Technol 2025; 59:103–118.

26. WHO. Guidelines for stopping mass drug administration and verifying elimination of human onchocerciasis. World Health Organisation, 2016: 1–55. Available at: https://www.who.int/publications/i/item/9789241510011. Accessed 20 December 2025.

27. Mitjà O, Gass K, Marks M, et al. Guidance for conducting and evaluating serological surveys to assess interruption of yaws transmission in the context of an eradication target. PLoS Negl Trop Dis 2025; 19:e0012899.

28. Kamau E, Gass K, Harding-Esch EM, et al. Design considerations for incorporating serological monitoring into trachoma prevalence surveys. medRxiv 2025; :2025.09.05.25334960.

29. Manz RA, Hauser AE, Hiepe F, Radbruch A. Maintenance of serum antibody levels. Annu Rev Immunol 2005; 23:367–386.

